# Cardiac Function Assessment with Deep-Learning-Based Automatic Segmentation of Free-Running 4D Whole-Heart CMR

**DOI:** 10.1101/2025.07.15.25331281

**Authors:** Augustin C. Ogier, Salomé Baup, Gorun Ilanjian, Aisha Touray, Angela Rocca, Jaume Banús, Isabel Montón Quesada, Martin Nicoletti, Jean-Baptiste Ledoux, Jonas Richiardi, Robert J. Holtackers, Jérôme Yerly, Matthias Stuber, Roger Hullin, David Rotzinger, Ruud B. van Heeswijk

## Abstract

**Background:** Free-running (FR) cardiac MRI enables free-breathing ECG-free fully dynamic 5D (3D spatial+cardiac+respiration dimensions) imaging but poses significant challenges for clinical integration due to the volume and complexity of image analysis. Existing segmentation methods are tailored to 2D cine or static 3D acquisitions and cannot leverage the unique spatial-temporal wealth of FR data.

**Purpose:** To develop and validate a deep learning (DL)-based segmentation framework for isotropic 3D+cardiac cycle FR cardiac MRI that enables accurate, fast, and clinically meaningful anatomical and functional analysis.

**Methods:** Free-running, contrast-free bSSFP acquisitions at 1.5T and contrast-enhanced GRE acquisitions at 3T were used to reconstruct motion-resolved 5D datasets. From these, the end-expiratory respiratory phase was retained to yield fully isotropic 4D datasets. Automatic propagation of a limited set of manual segmentations was used to segment the left and right ventricular blood pool (LVB, RVB) and left ventricular myocardium (LVM) on reformatted short-axis (SAX) end-systolic (ES) and end-diastolic (ED) images. These were used to train a 3D nnU-Net model. Validation was performed using geometric metrics (Dice similarity coefficient [DSC], relative volume difference [RVD]), clinical metrics (ED and ES volumes, ejection fraction [EF]), and physiological consistency metrics (systole–diastole LVM volume mismatch and LV–RV stroke volume agreement). To assess the robustness and flexibility of the approach, we evaluated multiple additional DL training configurations such as using 4D propagation-based data augmentation to incorporate all cardiac phases into training.

**Results:** The main proposed method achieved automatic segmentation within a minute, delivering high geometric accuracy and consistency (DSC: 0.94 ± 0.01 [LVB], 0.86 ± 0.02 [LVM], 0.92 ± 0.01 [RVB]; RVD: 2.7%, 5.8%, 4.5%). Clinical LV metrics showed excellent agreement (ICC > 0.98 for EDV/ESV/EF, bias < 2 mL for EDV/ESV, < 1% for EF), while RV metrics remained clinically reliable (ICC > 0.93 for EDV/ESV/EF, bias < 1 mL for EDV/ESV, < 1% for EF) but exhibited wider limits of agreement. Training on all cardiac phases improved temporal coherence, reducing LVM volume mismatch from 4.0% to 2.6%.

**Conclusion:** This study validates a DL-based method for fast and accurate segmentation of whole-heart free-running 4D cardiac MRI. Robust performance across diverse protocols and evaluation with complementary metrics that match state-of-the-art benchmarks supports its integration into clinical and research workflows, helping to overcome a key barrier to the broader adoption of free-running imaging.

## Introduction

Cardiovascular disease remains the leading cause of death globally,^1^ highlighting the critical need for accurate, efficient, and reproducible cardiac imaging tools to improve diagnosis. Cardiac magnetic resonance imaging (CMR) plays an important role in the assessment of cardiac anatomy and function, combining high soft-tissue contrast with a wide range of diagnostic parameters.^2^ However, conventional functional CMR protocols, particularly 2D multi-slice cine imaging, have limitations. These acquisitions usually require breath holding and are typically performed using thick slices (often 6–10 mm) and inter-slice gaps. Though they offer temporal coverage of the cardiac cycle, the poor through-plane resolution and dependence on pre-defined imaging planes may reduce their accuracy and reproducibility, especially in complex anatomies or when additional anatomical or functional information is requested after the scan has finished. Static 3D CMR acquisitions, such as whole-heart balanced steady-state free precession (bSSFP), overcome the limitations of slice thickness and provide a high (near-)isotropic spatial resolution, but they are generally acquired in a single cardiac phase (usually end-diastole) and are not meant to capture cardiac dynamics. Like 2D cine imaging, these acquisitions typically require ECG triggering and multiple breath-holds, which can be challenging in patients with limited breath-hold capacity or when the ECG signal quality is compromised. This dependence on patient cooperation and accurate timing adds complexity to the workflow and can impact image quality and reproducibility.

Free-running (FR) CMR has recently emerged as a powerful alternative for functional and anatomical cardiac imaging, offering motion-resolved, high-resolution 3D imaging of the entire heart across both cardiac and respiratory cycles.^3^ This technique combines a continuous free-breathing ECG-free acquisition with self-gated motion-resolved reconstruction, resulting in fully isotropic 5D datasets (isotropic 3D spatial + cardiac + respiratory dimensions). FR imaging enables retrospective reformatting in any orientation and provides both anatomical detail and functional coverage across all phases of the cardiac cycle. However, the ease with which FR datasets can be acquired stands in sharp contrast to the complexity of analyzing the vast number of images they generate. A single dataset can contain up to 70 short-axis slices across 4 respiratory and 25 cardiac phases – amounting to around 7,000 images per subject. This extensive information, while potentially clinically valuable, makes full manual segmentation unfeasible and currently prevents broader clinical adoption. As a result, prior validation and analyses of FR acquisitions have relied on careful reformatting of the isotropic volumes into a traditional stack of 2D short-axis (SAX) views to enable segmentation using conventional tools,^4–6^ ultimately negating the benefits of isotropic dynamic whole-heart imaging.

Deep learning (DL) methods have become the standard for automated CMR segmentation,^7^ and have been validated across cohorts such as the ACDC,^8^ UK Biobank,^9^ and MM-WHS.^10^ These benchmarks have played a central role in the development of fully automatic CMR segmentation methods, enabling systematic comparisons of architectures and serving as reference datasets for most segmentation studies in the field.^8,9,11–13^ While these efforts have led to major advances in automatic segmentation performance, they are inherently tied to classical CMR data formats. Most 2D cine segmentation models are trained on multi-slice SAX stacks with non-isotropic resolution.^8,9,11^ Segmentation of static 3D datasets provides high-resolution anatomical descriptions but is limited to a single cardiac phase and lacks dynamic information.^10,12,13^ Among existing dynamic true 3D CMR techniques comparable to FR imaging, 4D flow MRI offers a precedent for 4D segmentation.^14^ However, its emphasis on velocity encoding results in lower anatomical resolution and contrast, which limits its suitability for detailed structural segmentation.

To date, no DL method has been proposed to perform segmentation directly on the native isotropic 3D+cardiac dimension of FR CMR acquisitions.^7^ Existing segmentation frameworks are not readily transferable due to differences in image contrast, resolution, and dimensionality. Moreover, there is no publicly available benchmark dataset that includes manual annotations for FR imaging in its native form.

The goal of this study is therefore to develop and validate a dedicated automated segmentation framework for free-running CMR. The model is trained on a diverse cohort of healthy volunteers and heart failure patients, spanning multiple contrasts and magnetic field strengths. To overcome the impracticality of manual annotation across such large and dynamic datasets, we leverage a previously validated segmentation propagation approach based on diffeomorphic registrations^15^ to generate reference segmentation at end-diastole and end-systole. This approach enables fast high-resolution, full-cycle segmentation directly in the original 3D+cardiac space, without relying on 2D reformats. Crucially, we extend the traditional geometric-only evaluation by also assessing key clinical and physiological consistency metrics. By validating performance across both geometric and functional cardiac metrics, this work supports the broader clinical translation of 5D FR CMR and helps bridging the gap between advanced acquisition and clinical usefulness.

## Methods

### Study Population and Acquisition Protocol

All human studies were approved by the local ethics committee (CER-VD approvals 2022-00934 and 2021-02458), and written informed consent was obtained from all participants before scanning. Thirty-five subjects were prospectively scanned on two clinical MRI systems to assess the generalizability of the segmentation framework across field strength and contrast regimes (Figure 1.C). Dataset 1 (D_1_)^4^ included 15 healthy volunteers scanned at 1.5T (Magnetom Sola, Siemens Healthineers, Forchheim, Germany) with a FR acquisition based on a native (without injection of a contrast agent) bSSFP readout. Dataset 2 (D_2_)^16^ included 20 participants, including 10 healthy controls, 5 patients with heart failure with preserved ejection fraction (HFpEF), and 5 patients with heart failure with reduced ejection fraction (HFrEF), imaged at 3T (Magnetom PrismaFit, Siemens Healthineers) using an FR sequence based on a spoiled gradient-recalled echo (GRE) after a contrast agent injection (0.2 mmol/kg gadobutrol, Gadovist, Bayer AG, Leverkusen, Germany). The segmented 3D radial FR volumes were acquired in a canonical orientation (i.e. not oblique), centered on the left ventricle and with each consecutive radial interleave initiated by a line in the superior-inferior direction for the extraction of physiological motion signals.^17^ Demographic details are provided for both cohorts (Table 1). As a reference standard, all examinations also included a 2D cine protocol that was retrospectively ECG-gated and acquired during end-expiratory breath-holds. This compressed-sensing-accelerated protocol consisted of a stack of SAX cine plus single-slice 4-chamber and single-slice 2-chamber cines.^18^ Acquisition parameters are detailed for each pulse sequence and field strength (Table 2). All examinations were performed with the combination of a 32-channel spine coil and an 18-channel body coil.

**Figure 1:**
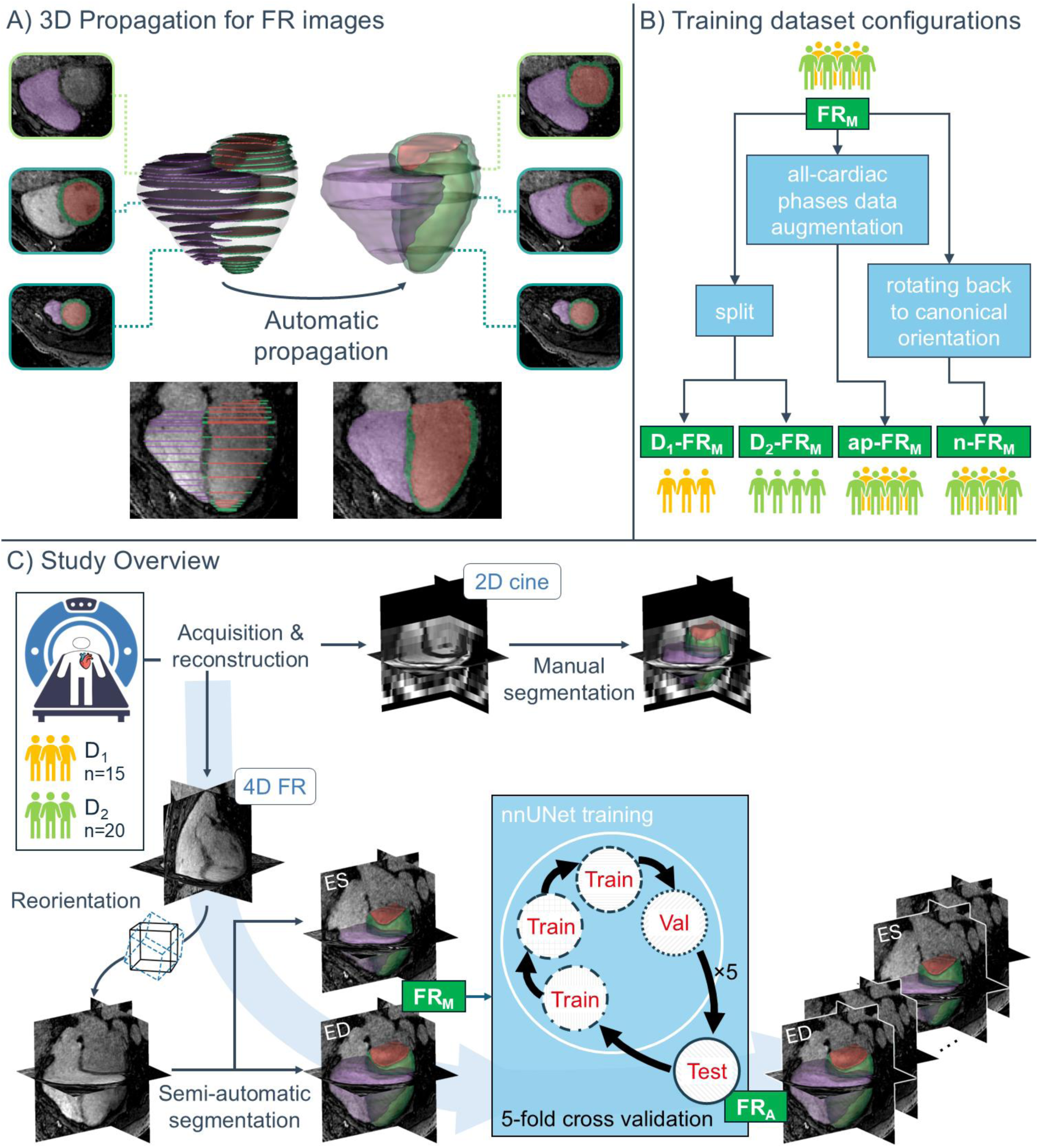
Workflow for semi-automatic and deep-learning segmentations of FR CMR. A) 3D-propagation scheme: Manual contours on a limited set of slices are extended to the entire 3D volume using a combination of diffeomorphic registrations. B) Training configurations: The main configuration (FRM) consists of semi-automatic segmentations on SAX-reformatted end-systolic (ES) and end-diastolic (ED) phases. Additional training configurations were derived to test specific hypotheses: cohort-specific training (D1-FRM, D2-FRM), 4D data augmentation including all cardiac phases (ap-FRM), and training in the native canonical space (n-FRM). C) Study design: Two cohorts (D1 at 1.5T; D2 at 3T) underwent 2D cine and FR imaging. Cine stacks were manually segmented, and FR volumes were reoriented and semi-automatically segmented at ED and ES via the 3D-propagation scheme in (A), yielding FRM. The main configuration FRM was used to train a nnU-Net model, yielding the automatic segmentation FRA under 5-fold cross-validation. All additional training configurations in (B) followed the same workflow.

**Table 1:**
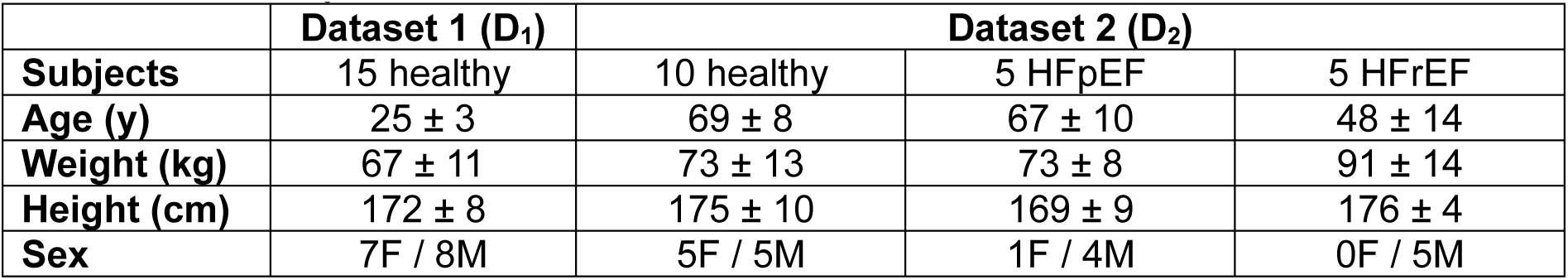
Participant demographics. HFpEF= heart failure with preserved ejection fraction, HFrEF= heart failure with reduced ejection fraction.

| | Dataset 1 ($D_1$) | Dataset 2 ($D_2$) | | |
| --- | --- | --- | --- | --- |
| **Subjects** | 15 healthy | 10 healthy | 5 HFpEF | 5 HFrEF |
| **Age (y)** | 25 ± 3 | 69 ± 8 | 67 ± 10 | 48 ± 14 |
| **Weight (kg)** | 67 ± 11 | 73 ± 13 | 73 ± 8 | 91 ± 14 |
| **Height (cm)** | 172 ± 8 | 175 ± 10 | 169 ± 9 | 176 ± 4 |
| **Sex** | 7F / 8M | 5F / 5M | 1F / 4M | 0F / 5M |

**Table 2:** MR image acquisition parameters for free-running (FR) and 2D cine acquisitions in datasets D_1_ and D_2_. Both cine and FR images were reconstructed using compressed sensing. The reported cine acquisition time includes full planning (from localizers to double-oblique plane prescription) and was dependent on patient breath-hold capacity.

| Sequence | 2D Cine |  | FR |  |
| --- | --- | --- | --- | --- |
| Dataset | $D_1$ | $D_2$ | $D_1$ | $D_2$ |
| Field strength | 1.5T | 3T | 1.5T | 3T |
| Sequence type | CS bSSFP | CS bSSFP | slab-selective bSSFP | slab-selective GRE |
| k-space trajectory | Undersampled Cartesian | Undersampled Cartesian | Segmented 3D radial spiral phyllotaxis | Segmented 3D radial spiral phyllotaxis |
| Dimensions | 2D | 2D | 3D | 3D |
| TE (ms) | 1.4 | 1.3 | 1.7 | 1.3 |
| TR (ms) | 3.2 | 3.4 | 3.5 | 3.0 |
| Flip angle (°) | 80 | 80 | Median 70 (range 58–70) | 15 |
| In-plane resolution (mm<sup>2</sup>) | 1.4 × 1.4 | 1.4 × 1.4 | 1.4 × 1.4 | 1.4 × 1.4 |
| **Slice thickness (mm)** | 8 | 8 | 1.4 — Isotropic slab | 1.4 — Isotropic slab |
| **Slice gap (mm)** | 2 | 2 | 0 | 0 |
| **Undersampling factor** | 7.5 | 8.6 | 2 – 3.3 Nyquist % per motion bin | 2 – 3.3 Nyquist % per motion bin |
| **Receiver bandwidth (Hz/pixel)** | 977 | 977 | 1116 | 1008 |
| **Field of view** | 360 × 284 mm<sup>2</sup> | 360 × 287 mm<sup>2</sup> | 220 × 220 × 220 mm<sup>3</sup> | 220 × 220 × 220 mm<sup>3</sup> |
| **ECG triggering** | Yes | Yes | No | No |
| **Respiratory motion handling** | Breath hold | Breath hold | Self-gated | Self-gated |
| **Acquisition time (min)** | ~ 10 | ~ 10 | 4:00 | 4:05 |

### 5D Image Reconstruction

For the free-running 5D images, cardiac and respiratory self-gating signals were extracted from the repeated superior–inferior projections of the segmented 3D radial trajectory.^3^ First, principal-component analysis generated several candidate respiratory signals. For each candidate component, Gaussian curves were fitted to every peak within the physiological breathing band of its power spectrum.^5^ A scoring metric, designed to favor components with a single dominant peak while down-weighting those with additional spurious peaks, was calculated, and the highest-scoring candidate was automatically selected as the respiratory trace.^5^ After subtracting this respiratory component, second-order blind identification isolated the dominant cardiac component,^19^ which was selected in the expected heart-rate range using the same spectral-ranking approach.

The validated motion components were used to retrospectively sort the continuous acquisition into four equally populated respiratory bins (end-expiration to end-inspiration) and non-overlapping 50 ms cardiac frames, yielding highly undersampled 5D k-space blocks. Compressed sensing (k-t sparse SENSE) was used to reconstruct 5D cardiac and respiratory motion-resolved images^3,20^, with regularization weights set to 0.001 for both temporal dimensions in D_1_, and 0.01 (cardiac) and 0.03 (respiratory) in D_2_. For the present study only the end-expiratory bins were retained, resulting in a 4D dataset (3D + cardiac cycle) per subject that spans the full cardiac cycle with minimal residual respiratory motion.

### Ground Truth Segmentation

Segmentation followed SCMR recommendations for biventricular analysis^21^ and produced segmentation masks for the left-ventricular blood pool (LVB), left-ventricular myocardium (LVM) and right-ventricular blood pool (RVB). The annotation protocol was further informed by prior literature aimed at standardizing ventricular segmentation.^22^ All contours were drawn and edited in FSLeyes.^23^ Although commercial CMR analysis software packages are commonly used in clinical practice, these platforms typically do not allow the exportation of segmentation masks for subsequent DL training, so they could not be employed for this study.

For the 2D cine reference data, end-diastolic (ED) and end-systolic (ES) frames were identified by visual inspection of the LVB area. Segmentation was performed slice-by-slice on the SAX cine, while the single-slice 4-chamber cine was consulted to determine the most basal SAX slice to include. A radiologist (with 2 years of experience) manually contoured the LVB and LVM, and a second radiologist (with 2 years of experience) delineated the RVB for both ED and ES phases.

For the FR volumes, the isotropic 4D images were rotated to conventional double-oblique cardiac axes by applying a rigid transform derived from the 2D cine SAX and 4-chamber orientations, allowing the subsequent manual work to follow SCMR guidelines that are defined for these views. After this transformation, the main imaging plane coincided with the anatomical short-axis view, whereas the two orthogonal planes corresponded approximately to the operator-prescribed long-axis orientations. Because a single global rigid transformation cannot always simultaneously match the exact angulation of both clinical short- and long-axis cines, these orthogonal planes were thus pseudo-2-chamber and pseudo-4-chamber views. ED and ES frames were then identified by visual inspection of the LVB area. Because each phase comprised 60–70 short-axis slices, fully manual segmentation was impractical. We therefore leveraged a previously validated semi-automatic segmentation pipeline.^15,24^ The same two radiologists who segmented the cine images manually contoured separate sets of key slices for ED and ES, including the most basal and apical planes, and added contours at fixed spatial steps or at levels showing significant anatomical change. After manual segmentation of the initial slices, the segmentations were automatically 3D-propagated independently for the ED and ES phases (Figure 1.A) through the remainder of the volume using a combination of diffeomorphic registration techniques,^15^ and the radiologists reviewed and performed minor local corrections on all propagated masks if needed. This semi-automatic workflow reduced per-subject annotation time while maintaining expert-level accuracy. The number of manually segmented and propagated slices was recorded for each subject.

Both the 2D cine and FR segmentations were subjected to two physiological consistency checks: conservation of myocardial volume between ED and ES and agreement of stroke volume between the left and right ventricles. To ensure myocardial volume conservation, LVM segmentations were adjusted as needed in an effort to keep the volume difference between ED and ES within a 10% tolerance. LV–RV stroke volume agreement was only assessed in healthy subjects, as regurgitation in patients could confound this metric. In cases of uncertainty, RVB segmentations, most often in 2D cine, were iteratively refined to aim for an RV stroke volume matching the LV stroke volume within a 10 mL tolerance. Any segmentation that failed either check was reviewed by a senior radiologist (12 years of experience), who applied targeted adjustments until both metrics fell within clinical tolerance, yielding the definitive ground-truth masks.

Hereafter, the fully manual 2D cine segmentations will be referred to as Cine_M_, and the semi-automatic FR segmentations as FR_M_. In order to quantify whether the rotation into the double-oblique orientation is needed for the subsequent DL training, we defined a second mask set, n-FR_M_ (for native-FR_M_), by applying the inverse of the cine-derived affine transform to FR_M_ (Figure 1.B), thereby transforming all contours into the canonical axial, sagittal, and coronal coordinate system of the original FR acquisition.

### Deep Learning Model and Training

For the DL model, we used the well-known nnU-Net^25^ enhanced with residual encoder framework, which has proven to be very competitive in several volumetric medical image segmentation challenges and has demonstrated strong performance across diverse tasks.^26^ We employed the 3D full-resolution configuration, as it provided the best results in preliminary testing. The model was trained using Dice loss, which is well-suited for medical image segmentation^27^ due to its direct optimization of spatial overlap between predicted and ground truth masks. The architecture, optimization parameters, and training schedule were fixed according to the default nnU-Net configuration.^25^ The batch size was set to 4 due to GPU memory limitations. The number of epochs was set to 80, based on empirical observation of training performance plateauing. The output consisted of 4 segmentation classes: background, LVB, LVM, and RVB. Trainings were run on a Linux workstation with two 24-core CPUs (Intel Xeon Gold 6248R; Intel, Santa Clara, CA, USA), 1.5 TB of RAM, and a 48Gb RTX A6000 GPU (NVIDIA Corporation, Santa Clara CA, USA). Both the total training time and the inference time were recorded.

To systematically validate our model across all subjects in the main configuration (FR_M_), we employed a 5-fold cross-validation strategy (Figure 1.C). The 35-subject cohort was divided into five non-overlapping folds of seven FR_M_ each (three D_1_ and four D_2_ subjects per fold). For each iteration, three folds (21 subjects, 42 three-dimensional volumes corresponding to ED and ES) were used for training, one fold (7 subjects, 14 three-dimensional volumes) for validation, and the remaining fold (7 subjects, 14 three-dimensional volumes) for testing, ensuring each fold served exactly three times as training, once as validation, and once as testing.

To evaluate the impact of data origin and data augmentation on segmentation performance, we implemented three additional training dataset configurations. These followed the same 5-fold logic but with adapted splits as needed (Figure 1.B). First, we trained one model exclusively on D_1_-FR_M_ subjects and another exclusively on D_2_-FR_M_ subjects, thereby isolating the effect of field strength and image contrast. In these configurations, cross-validation was limited to subjects within the corresponding cohort, with all subjects from the other cohort reassigned to the test set of the first fold to assess out-of-domain performance. Next, we trained a model using the back-transformed native-space labels n-FR_M_ to determine how effectively the network can learn directly in the unrotated canonical FR coordinate system without any cine-based reorientation. Finally, we devised a data-augmentation strategy by extending our 3D-propagation approach into a 4D-propagation method, propagating the semi-automatic 3D FR_M_ segmentations from ED and ES to all intermediate cardiac phases, creating an all-cardiac phases (ap) segmentation, which is hereafter denoted ap-FR_M_. Although this volume-to-volume propagation uses the same combination of diffeomorphic registration formalism, its application across whole 3D volumes, rather than across 2D slices, has never, and practically cannot, be validated against full 4D manual ground truth, since exhaustive annotation of every phase is prohibitive. Accordingly, 4D-propagation served solely as realistic data augmentation, drawing on prior work that uses non-linear registration to enrich volumetric training data^28^ and expanding our training set by roughly eightfold (≈16–20 volumes per subject instead of ED and ES only) without additional manual effort.

Upon inference completion, for all models, a post-processing step retained only the largest connected component per label, eliminating spurious islands of misclassification. Automatic segmentations produced by the models trained on FR_M_ are hereafter referred to as FR_A_; those from D_1_-FR_M_ as D_1_-FR_A_; from D_2_-FR_M_ as D_2_-FR_A_; from n-FR_M_ as n-FR_A_; and from ap-FR_M_ as ap-FR_A_.

### Validation Metrics

Validation of our 4D CMR processing framework relied on three complementary categories of metrics—geometric, clinical, and consistency—to fully characterize performance in both spatial and functional domains. Although the DL model automatically generates segmentations for every cardiac phase, all reported metrics were calculated only at ED and ES, since manual reference labels exist solely for those two phases.

To assess geometric accuracy, we compared paired segmentations defined in the same spatial frame (e.g. FR_M_ vs. FR_A_, D_1_-FR_M_ vs. D_1_-FR_A_, etc.) using established medical image segmentation metrics.^29^ Volumetric overlap was quantified by the Dice similarity coefficient (DSC), which measures the degree of spatial intersection between predicted and reference masks. Boundary agreement was characterized using the Hausdorff distance (HD), which is the maximum of the minimum distances between the predicted and ground-truth boundaries, as well as the average symmetric surface distance (ASSD), which reports the mean of those minimum boundary distances in both directions. We further evaluated systematic bias in volume estimation via the relative volume difference (RVD) defined as *RVD* = |*V*_*A*_ − *V*_*M*_| ⁄ *V*_*M*_, where *V*_*A*_ and *V*_*M*_ are the automatic and reference volumes, respectively. All geometric metrics were computed over the pooled set of ED and ES volumes rather than averaging phase-specific results, ensuring each voxel and each boundary discrepancy contributes equally to the overall assessment.

To evaluate clinical fidelity, we compared end-diastolic volume (EDV), end-systolic volume (ESV), and ejection fraction (EF), defined as *EF* = (*EDV* − *ESV*)⁄*EDV* × 100%, for both the left and right ventricular blood pools. Because these derived parameters are independent of image orientation or voxel grid, they could be computed and compared across any pair of segmentation sets (e.g. Cine_M_ vs. FR_M_ vs. FR_A_), providing a direct measure of cardiac anatomical and functional agreement.

To ensure physiological plausibility within each segmentation, we computed two consistency metrics. The left-ventricular myocardial volume mismatch *ɛ*_*LVM*_ was computed as the absolute difference between the ED and ES LVM volumes (EDV_LVM_ and ESV_LVM_, respectively) divided by the average of these two volumes, highlighting any non-physiological modification of myocardial mass from systole to diastole:

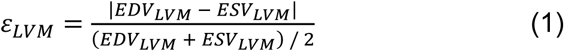

In addition, stroke volumes (SV = EDV − ESV) of the left (LVSV) and right (RVSV) ventricles were compared in healthy subjects only; agreement between LVSV and RVSV provided a second metric of intrinsic physiological validity.

All results are reported as mean ± standard deviation. Measurement agreement was evaluated by Bland–Altman analysis, providing bias and limits of agreement (LOA). Reproducibility and precision were quantified using the intra-class correlation coefficient with a (2, 1) formula^30^ and the corresponding standard error of measurement (SEM). Two-sided paired Student’s t-tests were used to compare normally distributed paired measurements (normality assessed with Shapiro–Wilk test). For *ɛ*_*LVM*_, a one-way repeated-measures ANOVA was performed to compare the different segmentation approaches, with post-hoc pairwise comparisons carried out using Tukey’s HSD procedure (with familywise α = 0.05).

## Results

For ground-truth generation, our 3D-propagation scheme reduced the need for manual contouring to 28 ± 5% of ED and ES slices. Specifically, at ED, 17 ± 5 RV and 18 ± 3 LV slices were manually segmented out of a total of 64 ± 7 RV and 68 ± 7 LV slices. At ES, 16 ± 4 RV and 17 ± 3 LV slices were contoured from total volumes of 50 ± 8 RV and 58 ± 8 LV slices. After propagation, fewer than 5% of slices required minor correction, adding, removing, or adjusting labels, to enforce myocardial volume conservation and stroke-volume agreement.

All DL trainings converged in 80 epochs (∼220 s per epoch, total ∼5 h), and all DL models consistently labeled cardiac structures across every cardiac phase (Supplementary Figure M1). Inference on a full 4D series required ∼1 min per subject. Only ED and ES frames were retained for subsequent analysis (Figure 2). Occasional larger segmentation failures occurred in cases with atypical anatomy or implanted devices, where basal slices or RV trabeculae were sometimes missegmented (Supplementary Figures S1–S2).

**Figure 2:**
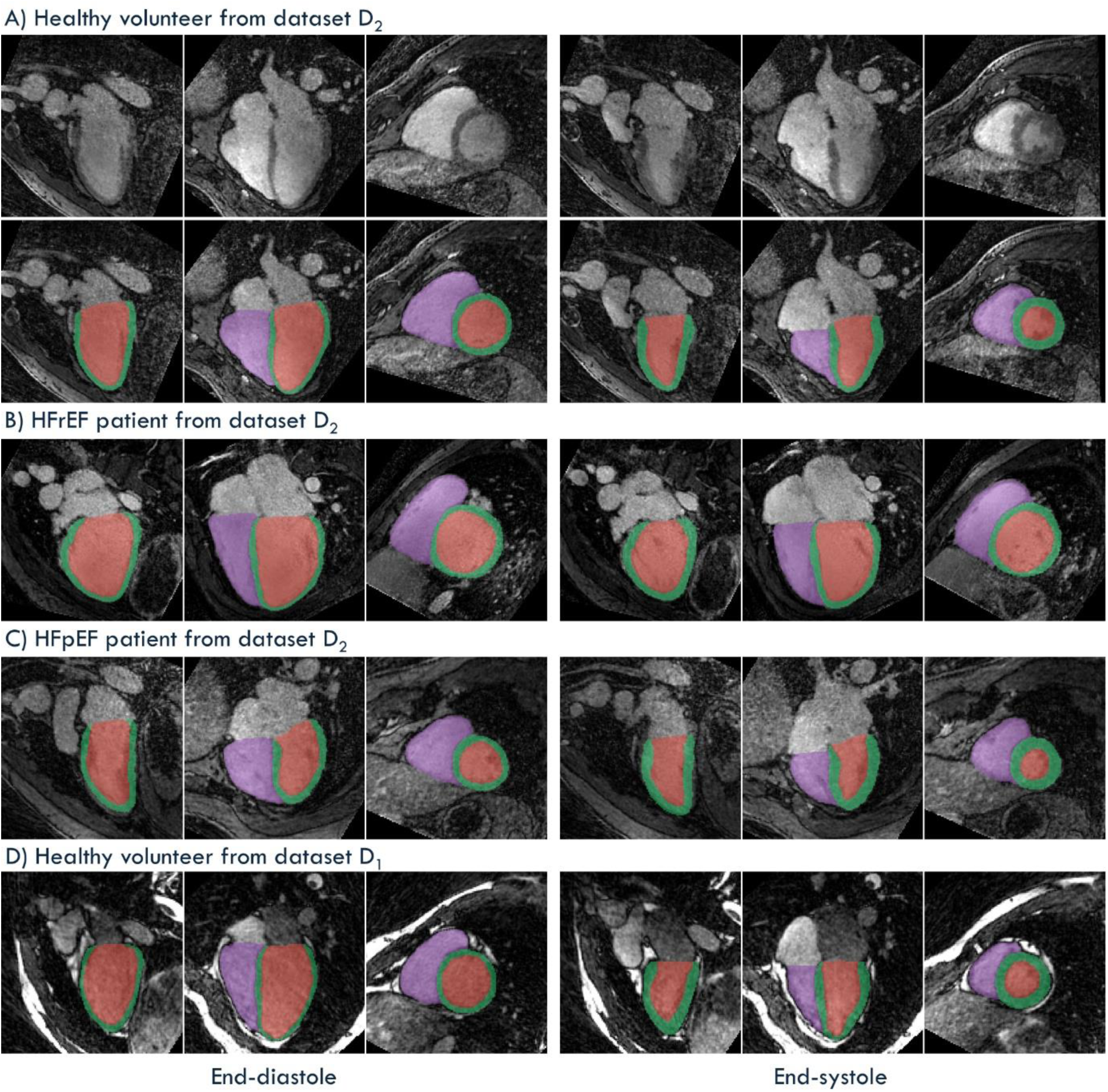
DL-based automatic segmentation (FR_A_) of spatially isotropic 4D free-running (FR) images at end-diastole (left) and end-systole (right). Examples are shown for A) a healthy volunteer in their 60s, B) a patient in their 30s with heart failure with reduced ejection fraction (HFrEF), and C) a patient in their 60s with heart failure with preserved ejection fraction (HFpEF), all scanned at 3T (dataset D_2_), as well as D) a healthy volunteer in their 20s scanned at 1.5T (dataset D_1_). The FR images are displayed in pseudo two-chamber, pseudo four-chamber, and short-axis views, with the three orthogonal planes corresponding to those used in the semi-automatic segmentation that served as ground truth for DL-based training. Segmented structures include the right ventricle blood pool (purple), left ventricle myocardium (green), and left ventricular blood pool (red).

### Geometric metrics

FR_A_ closely matched FR_M_ across all geometric metrics (Figure 3). The DSC between FR_A_ and FR_M_ was 0.94 ± 0.01 for LVB, 0.86 ± 0.02 for LVM, and 0.92 ± 0.01 for RVB. Boundary errors remained on the order of a single voxel for ASSD at 1.0 ± 0.1 mm (LVB), 0.9 ± 0.1 mm (LVM), and 1.2 ± 0.3 mm (RVB), while HD values were 6.2 ± 1.3 mm, 7.4 ± 2.6 mm, and 13.8 ± 6.5 mm, respectively. Manual-automatic volume bias was minimal, with RVD of 2.7 ± 2.1% for LVB, 5.8 ± 3.7% for LVM, and 4.5 ± 2.8% for RVB. The subject responsible for the HD outlier for RV segmentation is shown in Figure S1.

**Figure 3:**
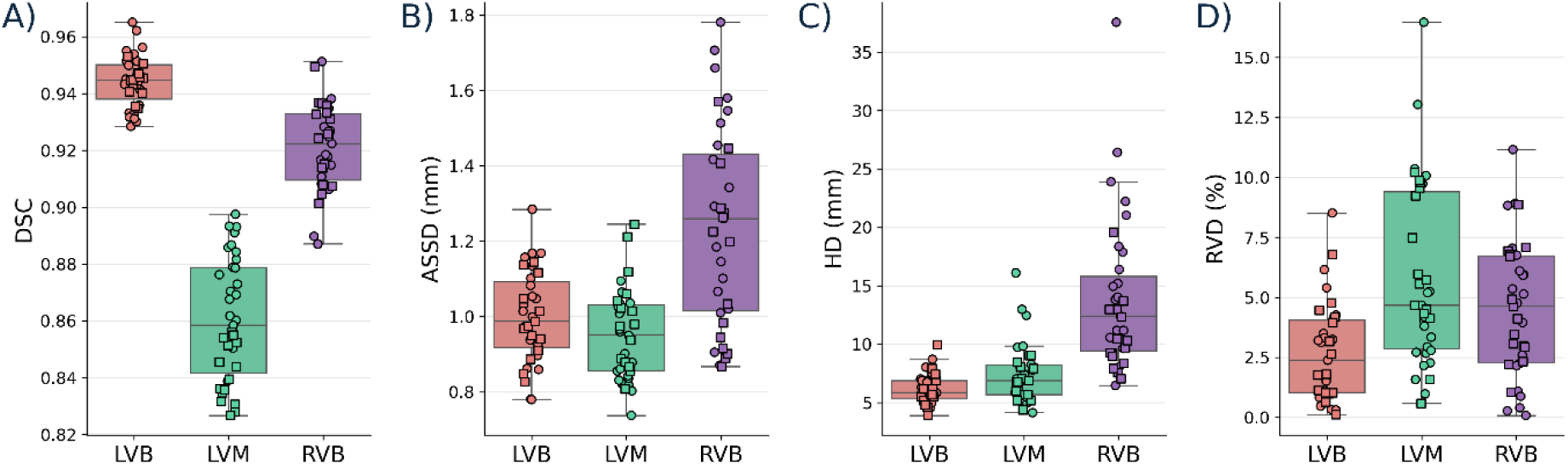
Geometric comparison between the ground truth segmentation (FR_M_) and the DL-based automatic segmentation (FR_A_) of the 4D FR images using metrics calculated from the combined end-diastolic and end-systolic frames. A) Dice similarity coefficients (DSC), B) average symmetric surface distance (ASSD), C) Hausdorff distance (HD), and D) relative volume difference (RVD) for the left ventricular blood pool (LVB), left ventricular myocardium (LVM), and right ventricular blood pool (RVB). Square markers represent subjects from dataset D_1_, and circle markers indicate subjects from dataset D_2_.

### Dataset-specific generalization

Models trained on all data (FR_A_) and on individual datasets (D_1_-FR_A_, D_2_-FR_A_) achieved similar high overlap and low volume bias when evaluated within their training cohort (Table 3). In contrast, cross-cohort performance dropped sharply for both overlap and bias: D_1_-FR_A_ on D_2_-FR_M_ yielded DSC < 0.65 and RVD > 32.0 %, while D_2_-FR_A_ on D_1_-FR_M_ resulted in DSC < 0.63 and RVD > 22.2 % for all regions of interest.

**Table 3:** Geometric accuracy and volume bias (DSC and RVD) for models trained on different datasets (D_1_, D_2_, and combined) and evaluated on FR_M_ segmentations from each cohort. FR_A_ denotes the model trained on the combined D_1_ and D_2_ cohort; D_1_-FR_A_ and D_2_-FR_A_ refer to models trained exclusively on D_1_ or D_2_, respectively. The combined model generalizes well across datasets, while single-dataset models perform best on their own cohort but poorly on the other. Metrics are reported as mean ± standard deviation.

| Trained on: | | $FR_A$ | | $D_1-FR_A$ | | $D_2-FR_A$ | |
| --- | --- | --- | --- | --- | --- | --- | --- |
| Validated against: | | $D_1-FR_M$ | $D_2-FR_M$ | $D_1-FR_M$ | $D_2-FR_M$ | $D_1-FR_M$ | $D_2-FR_M$ |
| DSC<br>(—) | LVB | $0.94 \pm 0.00$ | $0.94 \pm 0.01$ | $0.94 \pm 0.01$ | $0.65 \pm 0.24$ | $0.63 \pm 0.15$ | $0.94 \pm 0.01$ |
| | LVM | $0.84 \pm 0.01$ | $0.87 \pm 0.02$ | $0.83 \pm 0.01$ | $0.51 \pm 0.19$ | $0.43 \pm 0.16$ | $0.87 \pm 0.02$ |
| | RVB | $0.92 \pm 0.01$ | $0.92 \pm 0.02$ | $0.92 \pm 0.02$ | $0.59 \pm 0.2$ | $0.32 \pm 0.26$ | $0.92 \pm 0.02$ |
| RVD<br>(%) | LVB | $2.6 \pm 1.9$ | $2.7 \pm 2.1$ | $2.3 \pm 1.9$ | $32.0 \pm 27.9$ | $26.3 \pm 18.6$ | $3.1 \pm 2.1$ |
| | LVM | $6.2 \pm 3.0$ | $5.4 \pm 4.1$ | $6.0 \pm 3.6$ | $32.2 \pm 44.2$ | $22.2 \pm 21.9$ | $6.3 \pm 4.6$ |
| | RVB | $4.5 \pm 2.2$ | $4.5 \pm 3.1$ | $4.8 \pm 3.2$ | $48.8 \pm 48.2$ | $72.2 \pm 25.2$ | $5.4 \pm 3.6$ |

### Orientation and data augmentation effects

When applied to the native-space reference (n-FR_M_, the canonical orientation the data was acquired in), the reoriented model FR_A_ showed a slight decrease in both DSC and RVD (Table 4) indicating limited generalization of the models across rotations. However, training and evaluating directly in native space (n-FR_A_ on n-FR_M_) restored performance to levels equivalent to those achieved on reoriented volumes.

**Table 4:**
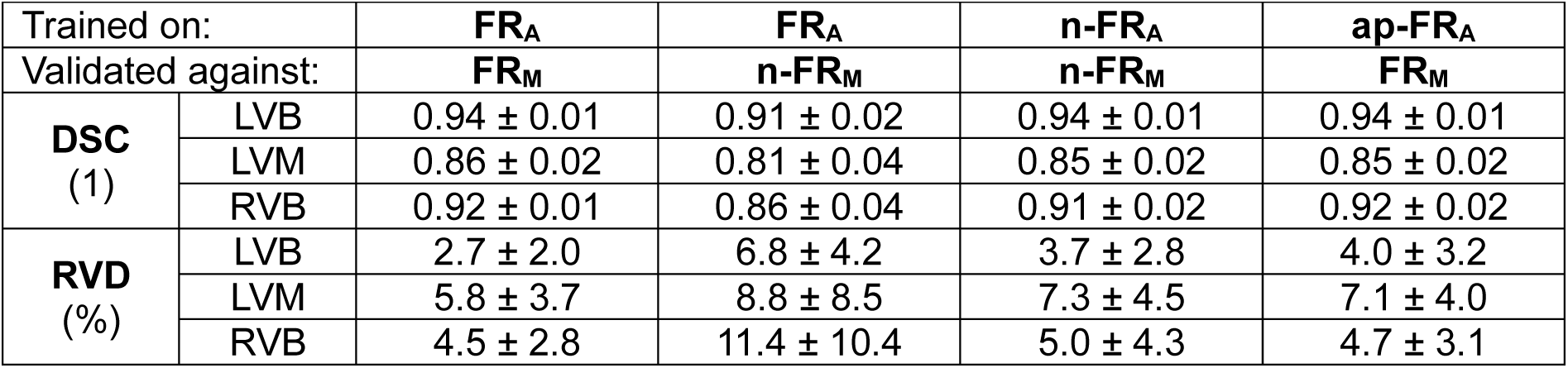
Geometric accuracy and volume bias (DSC and RVD) for different training configurations. Models were trained on native-space data (n-FR_A_), reoriented FR data (FR_A_), or FR_M_ with 4D propagation-based augmentation (ap-FR_A_), and validated against corresponding ground truth segmentations (n-FR_M_ or FR_M_). FR_A_ applied to native-space data showed a slight performance drop, while training directly in native space restored performance. The all-cardiac phases model ap-FR_A_ achieved comparable performance to FR_A_, indicating that the data augmentation strategy has a negligible effect on geometric accuracy. Metrics are reported as mean ± standard deviation.

| Trained on: |  | FR<sub>A</sub> | FR<sub>A</sub> | n-FR<sub>A</sub> | ap-FR<sub>A</sub> |
| --- | --- | --- | --- | --- | --- |
| Validated against: |  | FR<sub>M</sub> | n-FR<sub>M</sub> | n-FR<sub>M</sub> | FR<sub>M</sub> |
| **DSC**<br>(1) | LVB | 0.94 ± 0.01 | 0.91 ± 0.02 | 0.94 ± 0.01 | 0.94 ± 0.01 |
| | LVM | 0.86 ± 0.02 | 0.81 ± 0.04 | 0.85 ± 0.02 | 0.85 ± 0.02 |
| | RVB | 0.92 ± 0.01 | 0.86 ± 0.04 | 0.91 ± 0.02 | 0.92 ± 0.02 |
| **RVD**<br>(%) | LVB | 2.7 ± 2.0 | 6.8 ± 4.2 | 3.7 ± 2.8 | 4.0 ± 3.2 |
| | LVM | 5.8 ± 3.7 | 8.8 ± 8.5 | 7.3 ± 4.5 | 7.1 ± 4.0 |
| | RVB | 4.5 ± 2.8 | 11.4 ± 10.4 | 5.0 ± 4.3 | 4.7 ± 3.1 |

The all-cardiac phases model (ap-FR_A_) evaluated on FR_M_ resulted in high DSC and RVD that were very similar to FR_A_ on FR_M_ (Table 4), demonstrating that this data augmentation strategy has a negligible effect on geometric accuracy.

### Clinical metrics

Functional agreement between FR_M_ and FR_A_ was very high for the left ventricle (Figure 4). Bland-Atman analysis of EDV revealed very high reproducibility, though the small bias was statistically significant (p = 0.04), the LOA remained tight (–8.3; 12.0 mL). End-systolic volumes agreed even more closely (ICC = 1.0, SEM = 3.5 mL), and ejection fraction showed high concordance (ICC = 1.0, SEM = 2.0%, p = 0.3, LOA = –5.0; 5.9%). Right ventricular function followed a similar pattern, with high reproducibility (EDV ICC = 1.0, SEM = 6.4 mL; ESV ICC = 1.0, SEM = 7.4 mL; EF ICC = 0.9) and negligible bias, but wider LOA.

**Figure 4:**
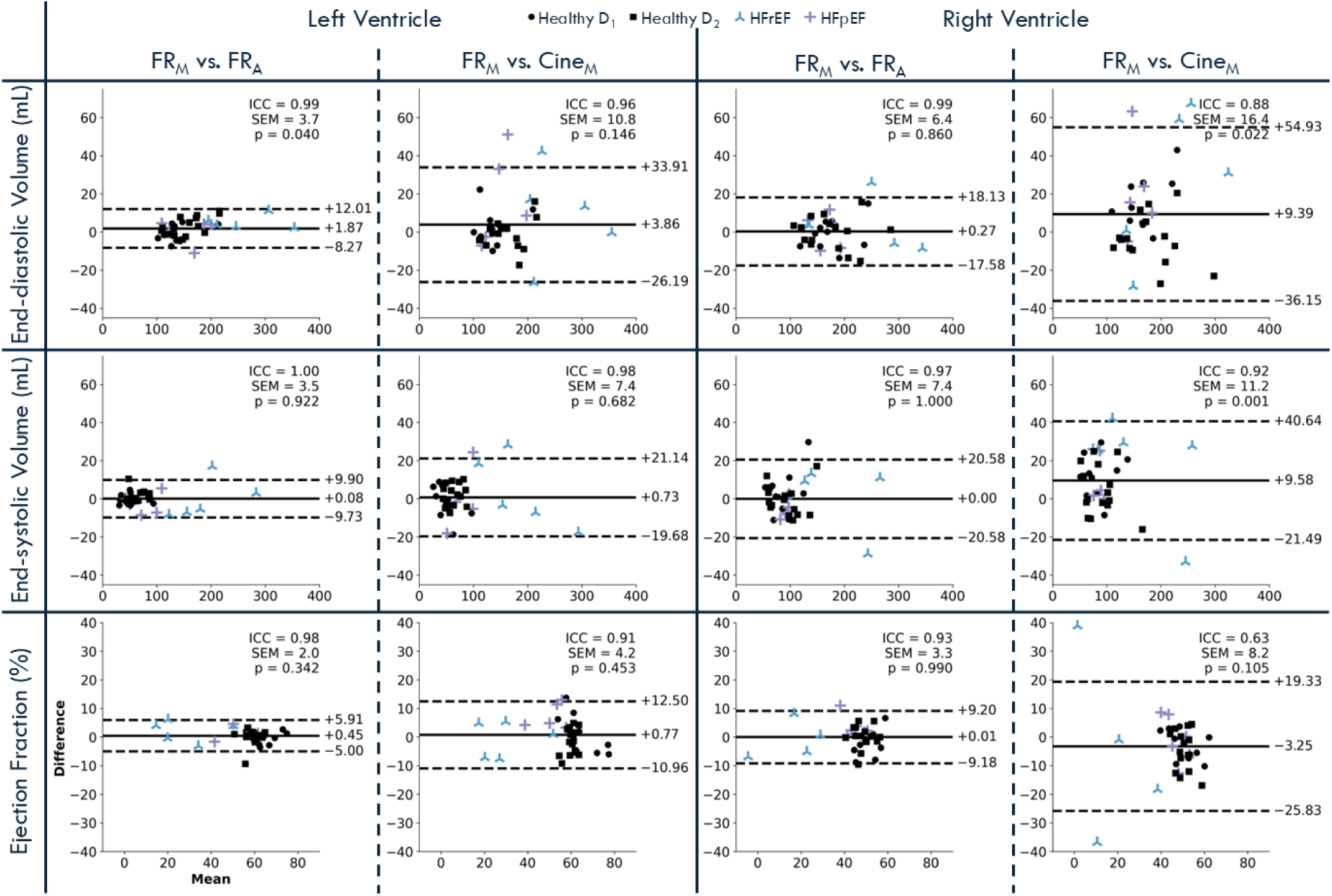
Clinical metrics comparison through volumetric and functional agreement. Bland-Altman analysis comparing semi-automatic segmentation (FR_M_) and deep learning-based automatic segmentation (FR_A_) of FR images, as well as FR_M_ and manual segmentation of cine images (Cine_M_), for end-diastolic volume, end-systolic volume, and ejection fraction in both the left and right ventricles. The x-axis represents the average of the two methods being compared, while the y-axis indicates the difference between them. The central solid line shows the bias, and dashed lines indicate the limits of agreement (bias ± 1.96 standard deviations). Intraclass correlation coefficients (ICC), standard error of measurement (SEM), and p-values for each comparison are also reported. Healthy volunteers are represented by black dots (square markers for dataset D_1_, circle markers for dataset D_2_), patients with HFrEF by blue crosses, and those with HFpEF by purple crosses.

In comparison, agreement between FR_M_ and Cine_M_ was high for the left ventricle, with ICCs of 0.96, 0.98, and 0.91 for EDV, ESV, and EF, respectively, but exhibited wider LOA. For the right ventricle, the LOA were even wider, and ICCs were lower (EDV ICC = 0.88, ESV ICC = 0.92, EF ICC = 0.66), reflecting reduced measurement reproducibility.

The clinical metrics for the n-FR_A_ and ap-FR_A_ configurations show that n-FR_A_ introduces only minimal degradation in reliability and accuracy compared to FR_A_ (with all ICC>0.90), whereas ap-FR_A_ exhibits a more pronounced, but still modest, increase (with all ICC>0.93) in LOA and bias (Supplementary Figure S3). For both configurations, the lowest reproducibility was observed for RV EF, although the bias remained below 1.5% with LOA around 10%.

### Physiological Consistency Metrics

The myocardial volume mismatch between systole and diastole *ɛ*_*LVM*_ (Figure 5) was the highest for Cine_M_ (7.1 ± 4.5%) followed by FR_M_ (5.6 ± 3.5%), with no significant difference between these two methods (p = 0.28). *ɛ*_*LVM*_ decreased non-significantly when moving from FR_M_ to FR_A_ (4.0 ± 2.4%; p = 0.24). n-FR_A_ (4.4 ± 3.4%) also did not differ from FR_A_ (p = 0.98). The lowest volume mismatch was observed for ap-FR_A_ (2.6 ± 1.9%), which was smaller than all other methods. Among automatic methods, it was the only one that differed significantly from FR_M_ (p =0.002).

**Figure 5:**
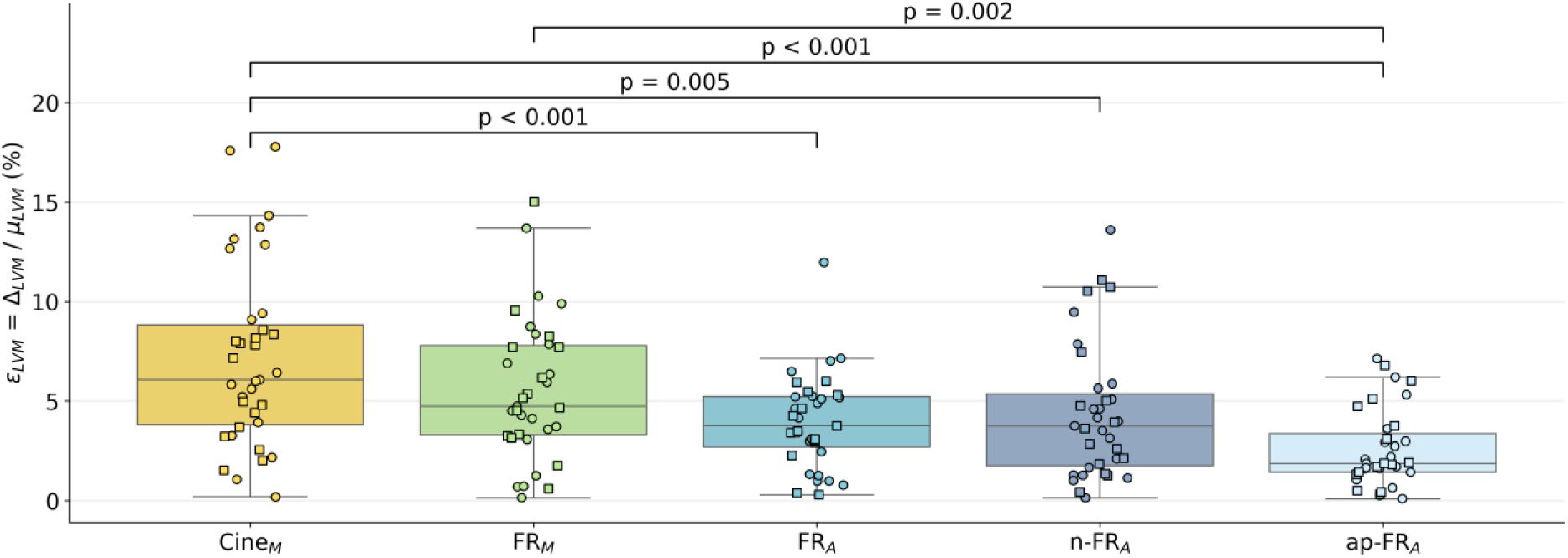
Systolic–diastolic myocardial volume mismatch. Left ventricular myocardium (LVM) volume mismatch *ɛ*_*LVM*_, defined as the difference between end-diastolic and end-systolic volumes (Δ_LVM_) divided by their mean (µ_LVM_), is shown for manual segmentation of cine images (Cine_M_), semi-automatic segmentation of 4D FR images (FR_M_), and three DL training dataset configurations (FR_A_, n-FR_A_, and ap-FR_A_). No significant difference was observed between Cine_M_ and FR_M_ (p = 0.28), while all FR-based models yielded significantly smaller errors than Cine_M_. Among automatic methods, ap-FR_A_ achieved the lowest *ɛ*_*LVM*_ and was the only one significantly different from FR_M_ (p = 0.002). Square markers represent subjects from dataset D_1_, and circle markers indicate subjects from dataset D_2_.

LV–RV stroke volume agreement remained high but exhibited progressively greater variability from Cine_M_ to FR_M_ to FR_A_ (Figure 6). Cine_M_ yielded an ICC of 0.94 with a modest bias of 2.8 mL (p = 0.04). Agreement for FR_M_ was similarly high (ICC = 0.91) but with increased bias (6.4 mL). FR_A_ showed wider LOA, a lower ICC of 0.83, and a bias of 7.1 mL, indicating that although all methods preserve stroke-volume parity, automatic segmentation introduces slightly greater variability. LV–RV stroke volume agreement for n-FR_A_ and ap-FR_A_ yielded ICCs, LOA, and biases nearly identical to FR_A_ (Supplementary Figure S4).

**Figure 6:**
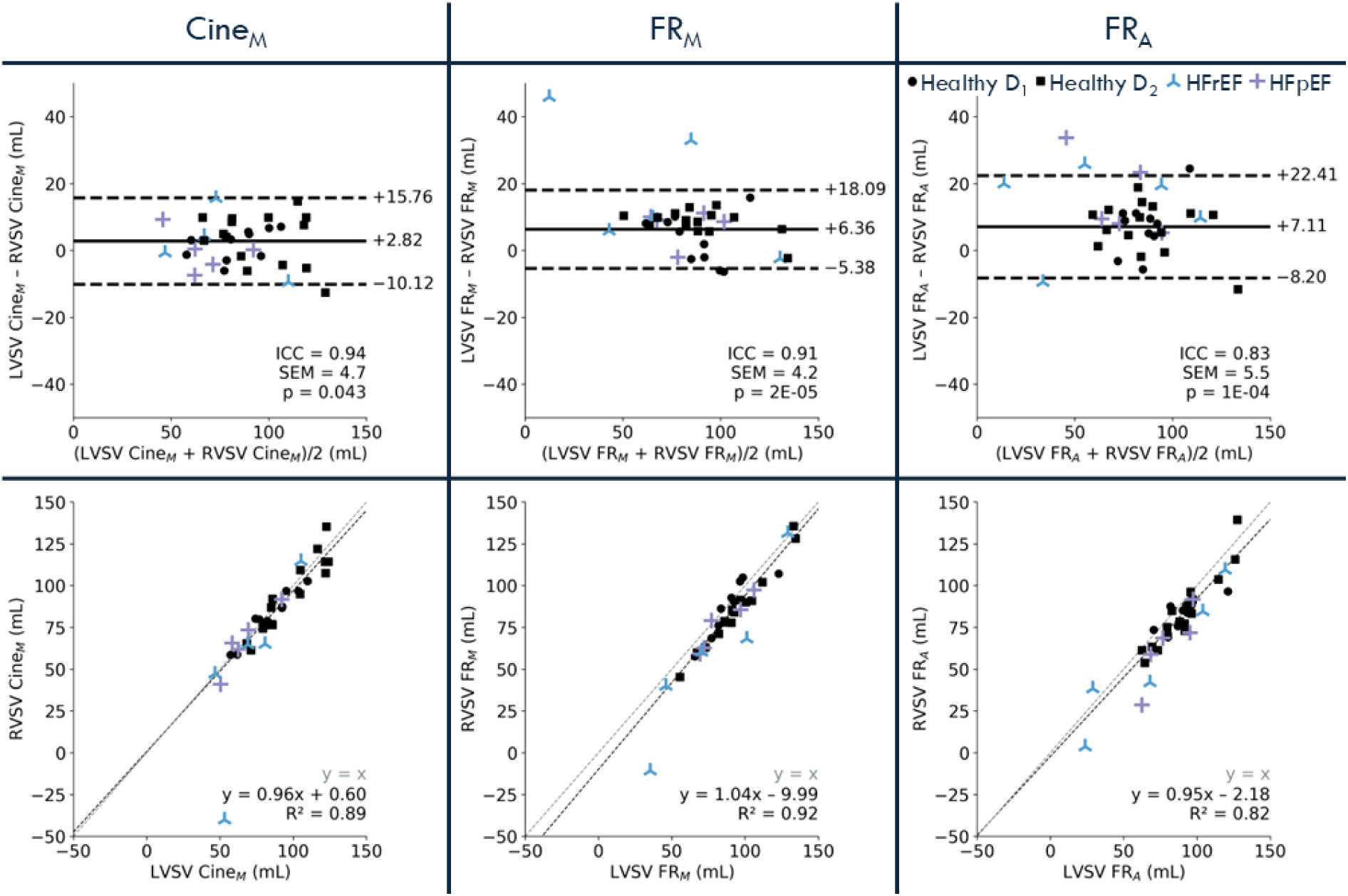
Stroke volume agreement. Bland-Altman analyses (top) and correlation plots (bottom) comparing stroke volumes of the right ventricle (RVSV) and left ventricle (LVSV), computed from manual segmentation of cine images (Cine_M_) and from semi-automatic (FR_M_) and deep learning-based automatic segmentation (FR_A_) of 4D FR images. In the Bland-Altman analysis, the central horizontal line represents the bias, and dashed lines indicate the limits of agreement (bias ± 1.96 SD). Agreement levels were calculated using only healthy volunteer data, since cardiac regurgitation was unknown for the patients. Intraclass correlation coefficients (ICC), standard error of measurement (SEM), and p-values for each comparison are also reported. Healthy volunteers are represented by black dots (square markers for dataset D_1_, circle markers for dataset D_2_), patients with HFrEF by blue crosses, and those with HFpEF by purple crosses.

## Discussion

In this study, we introduced and validated a deep-learning-based segmentation approach specifically tailored to isotropic dynamic (3D+t) FR cardiac MRI. Unlike existing segmentation methods designed for static 3D^10,12,13^ or multi-slice 2D^8,9,11^ acquisitions, this approach fully leverages the unique properties of FR imaging, including high isotropic spatial resolution, extensive spatial coverage, and comprehensive temporal dynamics without cine-based reformatting.

Our main method FR_A_ demonstrated excellent geometric accuracy across evaluated cardiac structures, achieving high DSC scores of 0.94 ± 0.01 (LVB), 0.86 ± 0.02 (LVM), 0.92 ± 0.02 (RVB). The corresponding HD values (6.2 ± 1.3 mm, 7.4 ± 2.6 mm, and 13.8 ± 6.5 mm) were in line with those reported in similar segmentation tasks.^8^ Limited volume bias was observed with RVD scores of 2.7 ± 2.0 (LVB), 5.7 ± 3.7 (LVM), and 4.5 ± 2.8 (RVB). Direct comparisons with prior studies are inherently complex due to the unique isotropic and dynamic nature of our FR datasets. Nonetheless, our achieved geometric metrics meet or exceed the current state-of-the-art reported for widely used CMR datasets. Bai et al. reported DSC scores of 0.94 for LVB, 0.88 for LVM, and 0.90 for RVB on the UK Biobank dataset,^9^ similar to our results, despite their images having lower through-plane resolution and inter-slice gaps. Using the ACDC challenge data, DSC values of 0.95 (LVB), 0.91 (LVM), and 0.92 (RVB) for top-performing methods were reported,^8,11^ though again with non-isotropic spatial resolution. Similarly, Bustamante et al. reported DSC values of 0.91 for LV and 0.89 for RV using 4D flow data,^31^ albeit with lower anatomical clarity and no myocardial segmentation due to limited contrast. On the MMWHS challenge, which includes static 3D whole-heart MR data, reported performances are generally lower for RVB and LVM: Muffoletto et al. achieved DSC scores of 0.87 (LVB), 0.69 (LVM), and 0.74 (RVB) using semi-supervised domain adaptation methods^12^, while Wang et al. reported values of 0.86 (LVB), 0.74 (LVM), and 0.85 (RVB) with a two-stage U-Net architecture.^13^ Notably, our method also exceeds the DSC values reported for intra-expert variability on the manual MMWHS annotations,^10^ although they remain slightly lower than the inter-expert variability reported in the ACDC challenge.^8^ Thus, despite the higher dimensional complexity, our method achieves comparable or higher accuracy, affirming its potential clinical relevance.

We extended validation beyond geometric overlap by directly assessing key clinical metrics. Comparing FR_A_ to FR_M_ for the LV, FR_A_ achieved an ICC > 0.99 for both EDV and ESV, with biases of 1.9 mL (LOA: –8.3; 12.0 mL) and 0.1 mL (–9.7; 9.9 mL), respectively. EF also showed excellent agreement (ICC = 0.98, bias = 0.45 %, LOA: –5.0; 5.9 %). These results closely mirror established intra-observer variability for 2D cine LV segmentation, where prior work^32^ reports an EDV bias of 1.9 mL (LOA: –7.9; 11.7 mL), an ESV bias of 0.6 mL (LOA: –6.7; 8.0 mL), and an EF bias of 0.2 % (LOA: –4.9; 4.6 %). For the RV, the clinical robustness of FR_A_ remained high (ICC: EDV = 0.98, ESV = 0.97, EF = 0.93) with minimal biases (0.27 mL, 0.00 mL, 0.01 %) but wider LOA (EDV: –17.6; 18.1 mL; ESV: –20.6; 20.6 mL; EF: –9.2; 9.2 %), reflecting the known challenges of RV delineation.

Consistent with previous studies,^7,32^ segmentation accuracy was notably higher for the LV than the RV. This discrepancy likely results from the manual segmentation process that is primarily performed in the short-axis orientation. This orientation is optimized for LV delineation and complicates precise RV segmentation, particularly at the basal RV region. The complexity of the manual RV segmentation impacts both the training and validation of the automatic method. Future RV segmentations may need to be performed in a dedicated oblique orientation.

We further introduced two physiological consistency metrics. We demonstrated lower myocardial volume mismatch between the ED and ES phases (*ɛ*_*LVM*_) in automatic segmentations compared to manual methods. Conversely, automatic segmentation slightly reduced LV–RV stroke volume agreement due to inherent challenges in RV segmentation accuracy. Notably, manual segmentations achieved consistency through iterative adjustments based on these metrics, simulating clinical practices, but this iterative process prolonged segmentation time. This underscores a key practical advantage of our automated method, which rapidly (∼1 minute per dataset) generates clinically relevant segmentations. These physiological consistency metrics do not rely on comparison to reference segmentations and could thus in the future serve to generate confidence scores for clinical use of our automatic segmentation method.

When trained on the combined D_1_ and D_2_ cohort, the FR_A_ model achieved high geometric accuracy compared to both D_1_-FR_M_ and D_2_-FR_M_, demonstrating effective learning across different contrasts and field strengths. In contrast, models trained exclusively on a single dataset (D_1_-FR_A_ or D_2_-FR_A_) performed well on their own cohort but showed significant accuracy drops when applied to the other, revealing limited cross-cohort transfer. Importantly, within each dataset, a model trained on that dataset alone matched the performance of the combined-training model on that same data. These results highlight that broad generalization requires diverse, multi-center, multi-field-strength, and multi-contrast FR datasets, as already explored by several recent studies,^5,6,33–35^ to ensure robust performance across varied clinical scenarios, whereas single-cohort models cannot reliably segment out-of-domain FR images.

Training the segmentation model in the native orientation (i.e. the canonical orientation it was acquired in) with n-FR_A_ effectively addressed the rotation sensitivity observed when applying the FR_A_ model trained on reoriented data back onto native volume datasets (n-FR_M_). By learning directly within the scanner’s native coordinate system rather than relying on cine-prescribed orientations, n-FR_A_ fully restored geometric accuracy. Specifically, n-FR_A_ on n-FR_M_ achieved DSC scores of 0.94 ± 0.01 (LVB), 0.85 ± 0.02 (LVM), and 0.91 ± 0.02 (RVB), with RVD scores of 3.7 ± 2.8%, 7.3 ± 4.5%, and 5.0 ± 4.3%, closely matching FR_A_ performance on rotated FR_M_ data. Agreement on clinical and physiological consistency metrics was also maintained. These results highlight the practical advantage of native-space training, streamlining future workflows by eliminating the need for supplementary acquisitions of oblique orientations, which can instead be performed post-hoc at the time of data analysis.

Augmenting the training dataset with 4D propagation-based data augmentation (ap-FR_A_) had minimal impact on per-phase accuracy and did not affect clinical accuracy. This evaluation is limited to ED and ES frames; ap-FR_A_ may improve segmentation accuracy on intermediate cardiac phases, though this remains unverified without all-cardiac phase ground truth segmentation. However, ap-FR_A_ notably improved temporal coherence, significantly reducing LVM volume mismatch *ɛ*_*LVM*_ from 4.0 ± 2.4% (FR_A_) to 2.6 ± 1.9%. This decrease underscores the potential value of 4D propagation in scenarios where temporal coherence across phases is particularly important, such as myocardial strain analyses, while remaining optional for standard volumetric or functional assessments.

An important contribution of this study is the semi-automatic generation of ground-truth segmentations, bridging the gap between fully manual annotations and the extensive datasets required for DL training. Although not fully manual, this semi-automatic approach relies on a validated method, making comprehensive annotation feasible for this study. The extent of segmentation required for accurate validation would have been impractical to achieve entirely manually. We publicly shared the code for our 3D and 4D propagation tools and FR reorientation script, alongside example datasets, via an open-access GitLab repository (gitlab.com/augustin-c-ogier/segpropa).

Although vision transformers^36^ and foundation models are gaining attention in medical imaging, we selected nnU-Net for its proven performance with relatively small datasets and its practicality. It consistently outperformed other methods in several challenges^26^ and adapts well to varied medical datasets without manual tuning. In contrast, transformer-based models require large-scale pretraining and remain largely untested on high-resolution 3D+t medical images. nnU-Net allowed efficient training from scratch on our FR data, making it a robust and accessible solution. Future work may explore transformers as annotated FR datasets grow.

Several limitations should nonetheless be recognized. Firstly, our segmentation included only LVB, LVM, and RVB. Comprehensive cardiac assessments would benefit from atrial segmentation, potentially enhancing ventricular delineation and valve boundary clarity. Secondly, while our model can be used to segment all cardiac phases, validation was limited to ED and ES frames due to the absence of full 4D ground truth annotations. A potential solution lies in further validating our 4D propagation method, which could serve as a reliable temporal reference and support the integration of advanced temporal architectures such as LSTM^37^. However, training on full 4D volumes would pose substantial memory and computational challenges. Likewise, our analysis focused solely on the end-expiratory phase of the reconstructed 5D data, and validation across the full 5D space, encompassing both cardiac and respiratory dynamics, remains methodologically complex, but a promising direction for future research.

Additionally, while tested across two magnetic field strengths and imaging contrasts, future work should assess performance across broader imaging conditions, including different contrast-enhanced protocols,^33,34^ low-field systems,^6^ more varied patients cohorts, and other acquisition variants using modified pulse sequences.^5,35^ Although the model performed well across a range of patient cases, it showed reduced accuracy in subjects with markedly atypical anatomy. Future work is needed to assess whether such approaches remain robust in populations with anatomical variability, such as patients with congenital heart disease.^34^

In conclusion, free-running imaging offers substantial advantages over conventional 2D cine MRI, including improved easy-of-use and time efficiency, isotropic spatial resolution, and rich spatiotemporal information. However, the analytical complexity of processing 5D data has long posed a barrier to clinical adoption. By introducing an accurate, rapid, and scalable segmentation framework that helps mitigate this barrier. Notably, our validation extends beyond standard geometric metrics to include clinically meaningful indices and physiological consistency measures, offering a more complete evaluation than most segmentation studies to date. This comprehensive assessment supports the reliability of our approach not only for anatomical delineation but also for functional analysis. By enabling efficient analysis of dynamic whole-heart data, this work may contribute to the integration of FR CMR into clinical workflows with limited cardiac MR expertise.

## Abbreviations

2D: two-dimensional
3D: three-dimensional
4D: four-dimensional
ACDC: Automated Cardiac Diagnosis Challenge
ASSD: average symmetric surface distance
bSSFP: balanced steady-state free precession
CS: compressed sensing
CMR: cardiovascular magnetic resonance
DL: deep learning
DSC: Dice similarity coefficient
ED: end-diastole
EDV: end-diastolic volume
EF: ejection fraction
ES: end-systole
ESV: end-systolic volume
GRE: gradient-recalled echo
HD: Hausdorff distance
HFpEF: heart failure with preserved ejection fraction
HFrEF: heart failure with reduced ejection fraction
ICC: intra-class correlation coefficient
LSTM: long short-term memory
MMWHS: multi-modality whole-heart segmentation
PCA: principal-component analysis
RV: right ventricle/right ventricular
RVD: relative volume difference
SAX: short-axis
SCMR: Society for Cardiovascular Magnetic Resonance
SEM: standard error of measurement
TR: repetition time
TE: echo time
LOA: limits of agreement

## Statements

## Data and code availability

The code used for both the 3D and 4D propagation methods, as well as the reorientation of free-running images to match 2D cine views, is available on GitLab at *gitlab.com/augustin-c-ogier/segpropa*, along with a representative 3T gadolinium-based contrast agent (GBCA)– enhanced dataset for testing and demonstration purposes [the latter upon acceptance of the manuscript]. The nnU-Net framework is available from the original authors at *github.com/MIC-DKFZ/nnUNet*. The majority of datasets cannot be shared due to restrictions imposed by local ethics regulations. Consequently, the trained deep learning models, which rely on this data, are also not publicly available. However, both can be made available upon reasonable request and pending appropriate approvals.

## Regulatory approval

Ethics approval was obtained from the Ethics Committee of the Canton of Vaud (CER-VD) of Switzerland under reference numbers 2022-00934 and 2021-02458. All participants provided written informed consent to participate prior to enrollment.

## Supplementary Figures

**Supplementary Figure S1:**
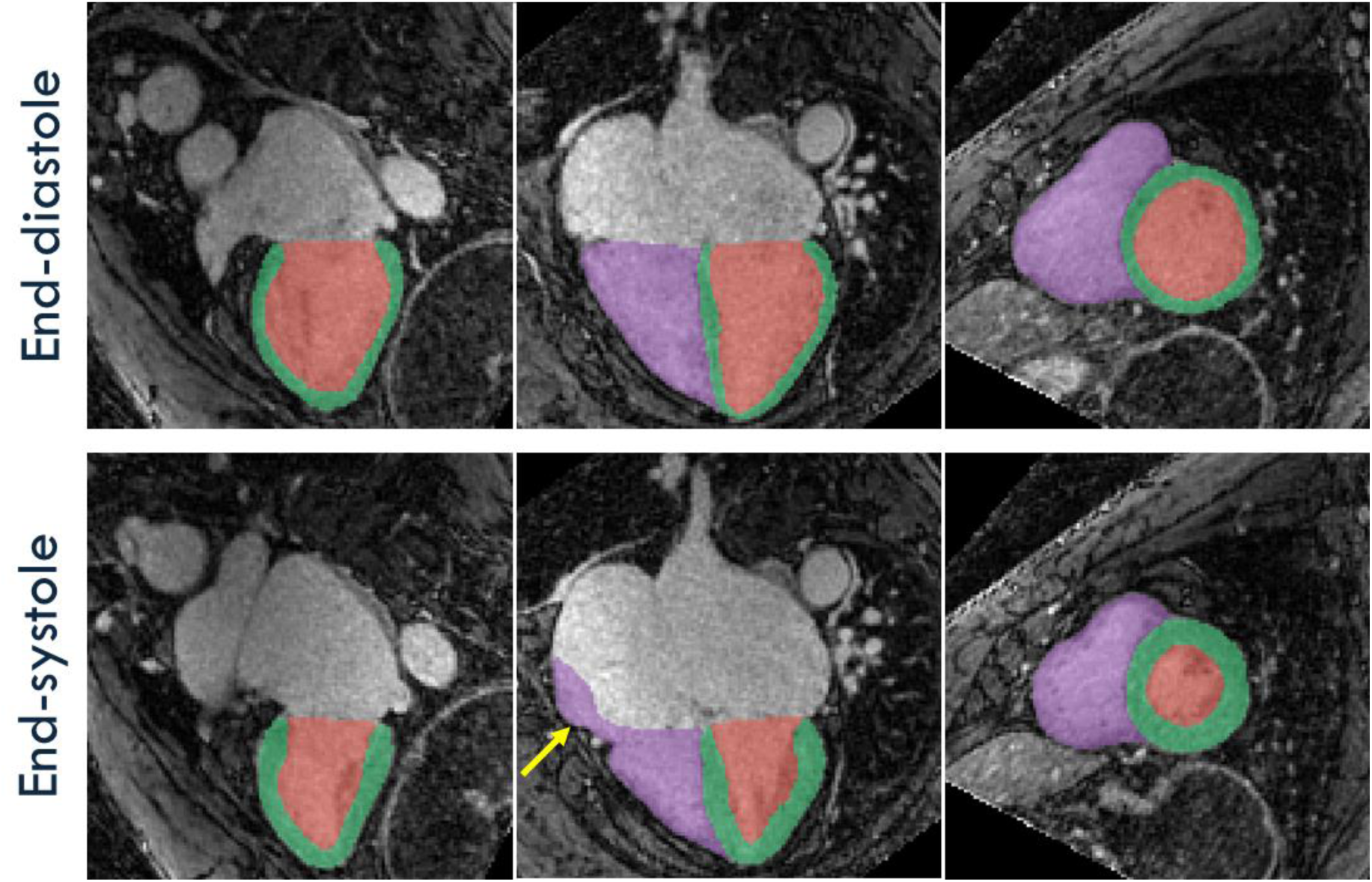
Segmentation mismatch in the case of non-standard anatomy. DL-based automatic segmentation (FR_A_) of spatially isotropic 4D free-running (FR) images at end-diastole (top) and end-systole (bottom) for a patient in their 70s with HFpEF, acquired at 3T (dataset D_2_). The subject presents with marked bi-atrial dilation. FR images are shown in pseudo two-chamber, pseudo four-chamber, and short-axis view (left to right), corresponding to the orthogonal planes used during semi-automatic ground truth segmentation. The right ventricle blood pool (purple), the left ventricle myocardium (green), and left ventricle blood pool (red) are highlighted. The automatic segmentation shows an overextension at end-systole only of the right ventricle contour into the right atrium (arrow), illustrating a common challenge in RV delineation under pathological remodeling.

**Supplementary Figure S2:**
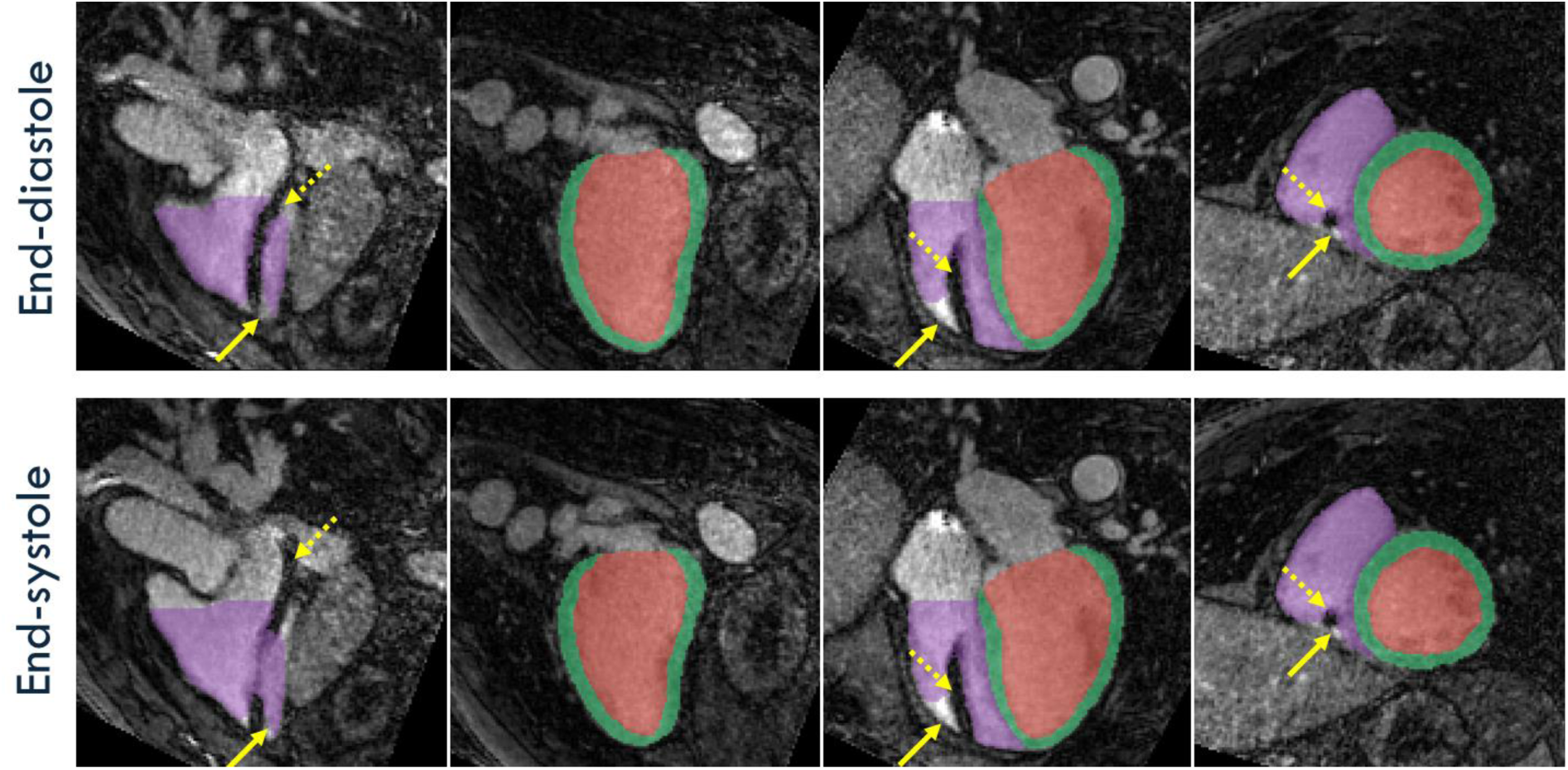
Partial segmentation failure in the presence of a device artifact. DL-based automatic segmentation (FR_A_) of spatially isotropic 4D free-running (FR) images at end-diastole (top) and end-systole (bottom) for a patient in their 60s with HFrEF, acquired at 3T (dataset D_2_), showing an imaging artifact (dashed arrows) due to a implantable cardioverter defibrillator (ICD) lead (Acticor 7 VR-T, Biotronik, Berlin, Germany) in the right ventricle (RV). FR images are shown in RV pseudo two-chamber, LV pseudo two-chamber, pseudo four-chamber, and short-axis view (left to right). The DL model segmented the RV blood pool (purple), left ventricle myocardium (green), and left ventricle blood pool (red) accurately across the heart, except in the immediate vicinity of the ICD artifact (dashed arrows), where segmentation performance was reduced (solid arrows). This case highlights the model’s general robustness, along with challenges posed by uncommon artifacts.

**Supplementary Figure S3.**
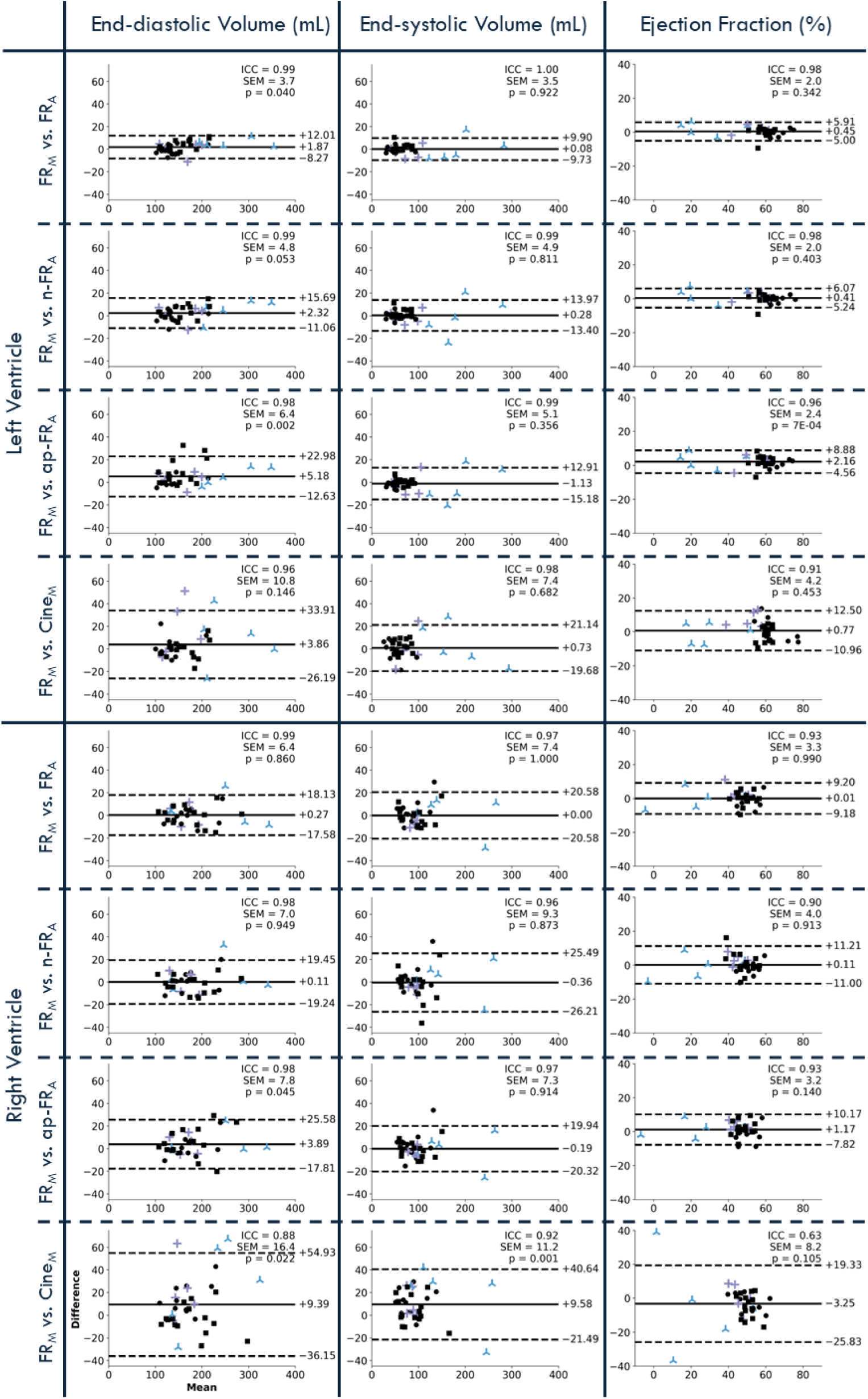
Clinical metrics comparison through volumetric and functional agreement for all segmentation strategies. Bland-Altman analysis comparing semi-automatic segmentation (FR_M_) with four other methods—manual segmentation of cine images (Cine_M_), standard deep learning-based segmentation (FR_A_), native-space model (n-FR_A_), and all cardiac phases (ap-FR_A_)—for end-diastolic volume, end-systolic volume, and ejection fraction in both the left and right ventricles. Healthy volunteers are represented by black dots (square markers for dataset D_1_, circle markers for dataset D_2_), patients with HFrEF by blue crosses, and those with HFpEF by purple crosses.

**Supplementary Figure S4:**
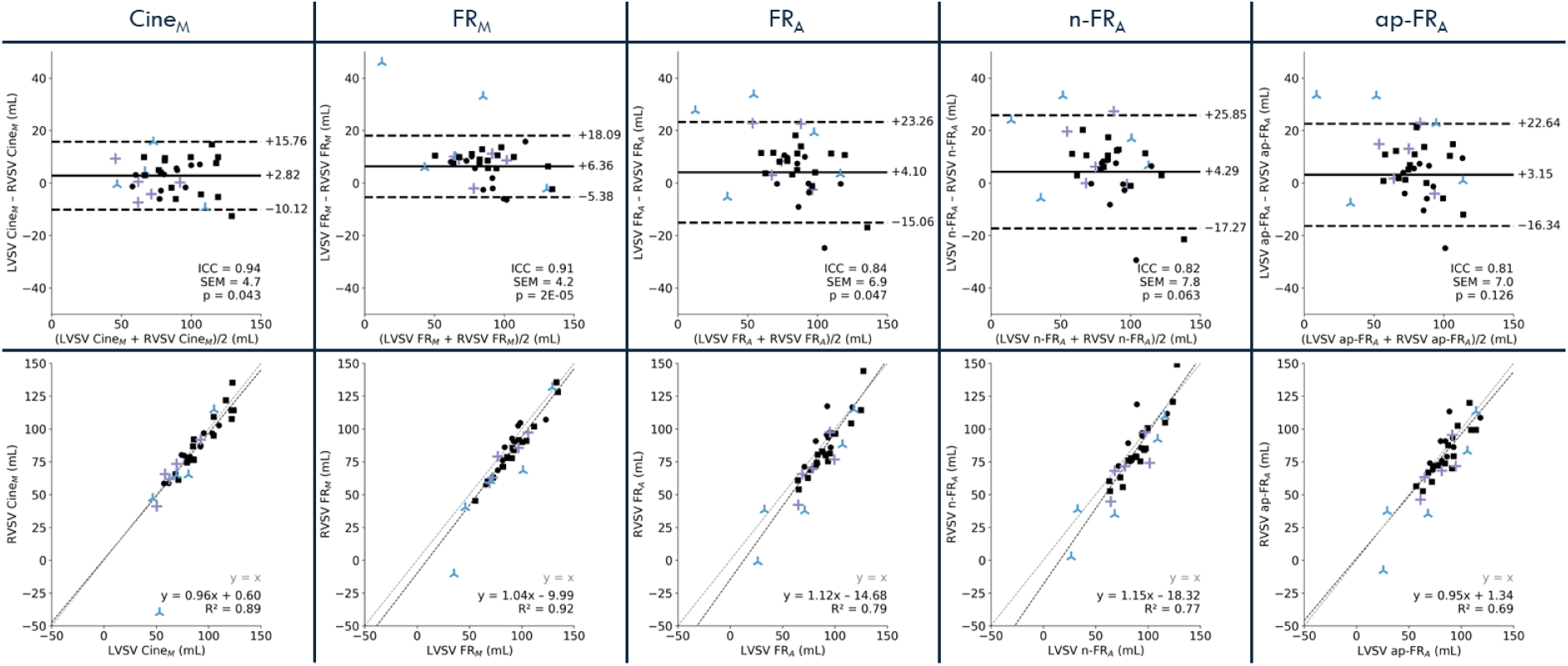
LV–RV stroke volume agreement. Bland-Altman analysis (top) and correlation plots (bottom) comparing stroke volumes of the right ventricle (RVSV) and left ventricle (LVSV), computed from manual segmentation of cine images (Cine_M_) and from semi-automatic segmentation of 4D FR images (FR_M_), and automatic deep learning-based segmentations (FR_A_, n-FR_A_, and ap-FR_A_). Healthy volunteers are represented by black dots (square markers for dataset D_1_, circle markers for dataset D_2_), patients with HFrEF by blue crosses, and those with HFpEF by purple crosses.

Supplementary Figure M1: A) 3D visualization of the heart region throughout the cardiac cycle, showing three orthogonal planes and dynamic surface meshes of the left ventricular blood pool (LVB, red), left ventricular myocardium (LVM, green), and right ventricular blood pool (RVB, purple). B) Free-running (FR) images are displayed in pseudo two-chamber, pseudo four-chamber, and short-axis views (left to right), corresponding to the orthogonal planes used in the semi-automatic segmentation that served as ground truth for deep learning training. Segmented structures are color-coded as in the 3D view. For both A and B, the frame rate is 50 ms, matching the temporal resolution of the reconstructed images. Note that while the slice orientations are consistent between A and B, the slice positions are not spatially matched. Data shown is from a healthy volunteer in their 60s, acquired at 3T (dataset D_2_).

