## Supplementary figures and images for "Cardiac Function Assessment with Deep-Learning-Based Automatic Segmentation of Free-Running 4D Whole-Heart CMR"

### Figure M1

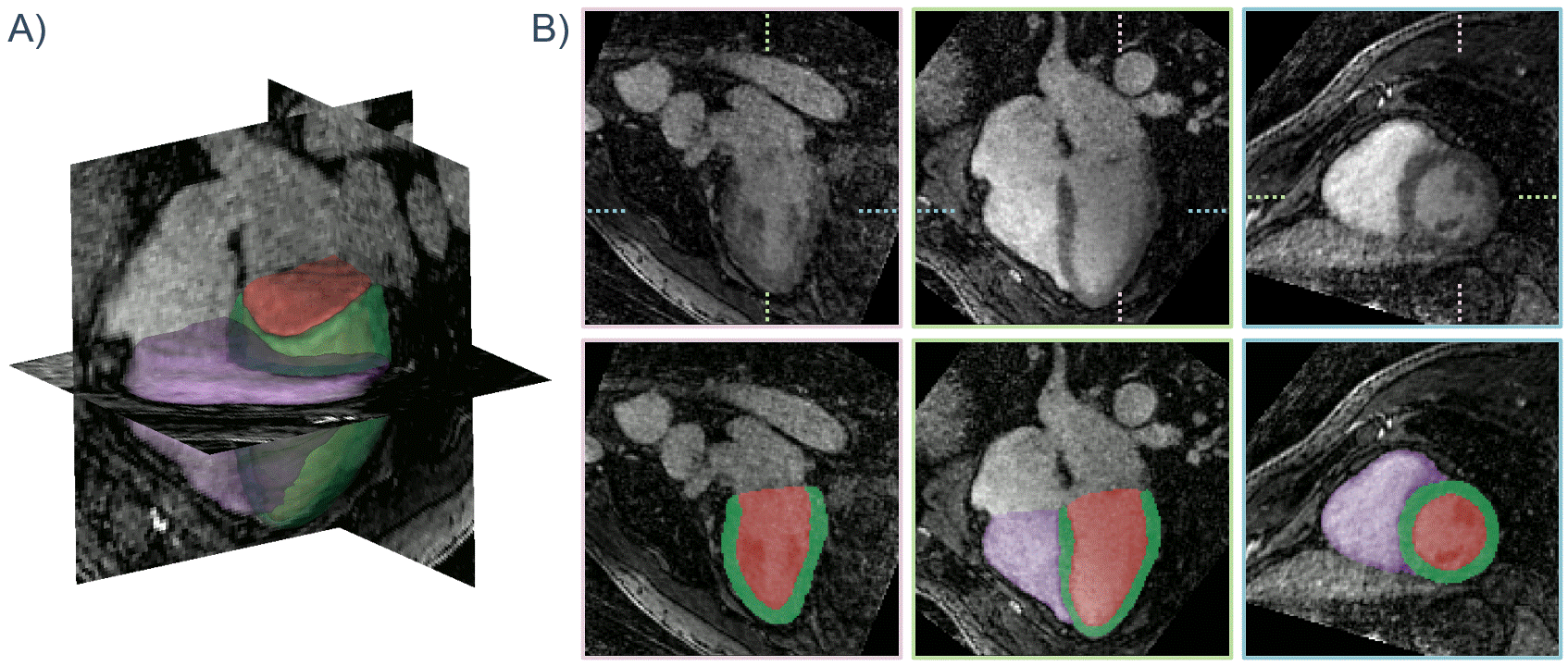
